# Integrin/TGF-β1 inhibitor GLPG-0187 blocks SARS-CoV-2 Delta and Omicron pseudovirus infection of airway epithelial cells which could attenuate disease severity

**DOI:** 10.1101/2022.01.02.22268641

**Authors:** Kelsey E. Huntington, Lindsey Carlsen, Eui-Young So, Matthias Piesche, Olin Liang, Wafik S. El-Deiry

## Abstract

As COVID-19 continues to pose major risk for vulnerable populations including the elderly, immunocompromised, patients with cancer, and those with contraindications to vaccination, novel treatment strategies are urgently needed. SARS-CoV-2 infects target cells via RGD-binding integrins either independently or as a co-receptor with surface receptor angiotensin-converting enzyme 2 (ACE2). We used pan-integrin inhibitor GLPG-0187 to demonstrate blockade of SARS-CoV-2 pseudovirus infection of target cells. Omicron pseudovirus infected normal human small airway epithelial (HSAE) cells significantly less than D614G or Delta variant pseudovirus, and GLPG-0187 effectively blocked SARS-CoV-2 pseudovirus infection in a dose-dependent manner across multiple viral variants. GLPG-0187 inhibited Omicron and Delta pseudovirus infection of HSAE cells more significantly than other variants. Pre-treatment of HSAE cells with MEK inhibitor (MEKi) VS-6766 enhanced inhibition of pseudovirus infection by GLPG-0187. Because integrins activate TGF-β signaling, we compared plasma levels of active and total TGF-β in COVID-19+ patients. Plasma TGF-β1 levels correlated with age, race, and number of medications upon presentation with COVID-19, but not with sex. Total plasma TGF-β1 levels correlated with activated TGF-β1 levels. In our preclinical studies, Omicron infects lower airway lung cells less efficiently than other COVID-19 variants. Moreover, inhibition of integrin signaling prevents SARS-CoV-2 Delta and Omicron pseudovirus infectivity, and may mitigate COVID-19 severity through decreased TGF-β1 activation. This therapeutic strategy may be further explored through clinical testing in vulnerable and unvaccinated populations.

## Introduction

Although several highly effective vaccines have now been developed against coronavirus disease 2019 (COVID-19), its threat to public health persists due to the presence of breakthrough cases, the current improbability of achieving herd immunity, reluctance to vaccinate among significant segments of the population, less available vaccines in much of the developing world, and the emergence of highly transmissible and immune evasive Delta and Omicron variants. Therefore, novel treatment strategies are urgently required to prevent severe disease, hospitalization and death, especially in vulnerable populations such as the elderly and immunocompromised, as well as those with pre-existing conditions including patients with cancer, or who cannot get vaccinated. Here, we present integrin inhibition with or without MEK inhibition as a potential treatment strategy for severe COVID-19.

Severe acute respiratory syndrome coronavirus 2 (SARS-CoV-2), the virus responsible for COVID-19, enters cells via interaction of its spike protein with angiotensin-converting enzyme 2 (ACE2), and possibly other receptors such as CD147/26, on human cells (1-4). The receptor-binding domain of spike contains upstream from the ACE2 binding site a novel RGD (Arg-Gly-Asp) motif that is absent in SARS-1. The RGD motif was originally identified within several extracellular matrix proteins as the minimal peptide sequence required for cell attachment via integrins (5-7), which make transmembrane connections to the cytoskeleton and activate many intracellular signaling pathways (8). Integrins are also commonly used as receptors by many human viruses (9). The conservation of the motif and its localization in the receptor-binding region of the SARS-CoV-2 spike protein suggests that integrins may serve as alternative receptors for this virus (10). Indeed, recent evidence suggests that this motif allows SARS-CoV-2 binding to integrins on human cells (11), facilitating viral infection (12) **(Figure 1)**, which may contribute to the higher transmission efficiency compared to SARS-1. There are eight known RGD-binding integrins with potential to impact on the pathogenesis of SARS-CoV-2: αvβ1, αvβ3, αvβ5, αvβ6, αvβ8, α5β1, α8β1, and αIIbβ3. A recent study has shown that blocking integrin αVβ3 prevents SARS-CoV-2 from binding to the vascular endothelium, potentially inhibiting virus-induced loss of endothelial barrier integrity and spread of SARS-CoV-2 to other organs (13). Thus, therapeutic inhibition of RGD-binding integrins may provide benefit to COVID-19 patients.

**Figure 1.**
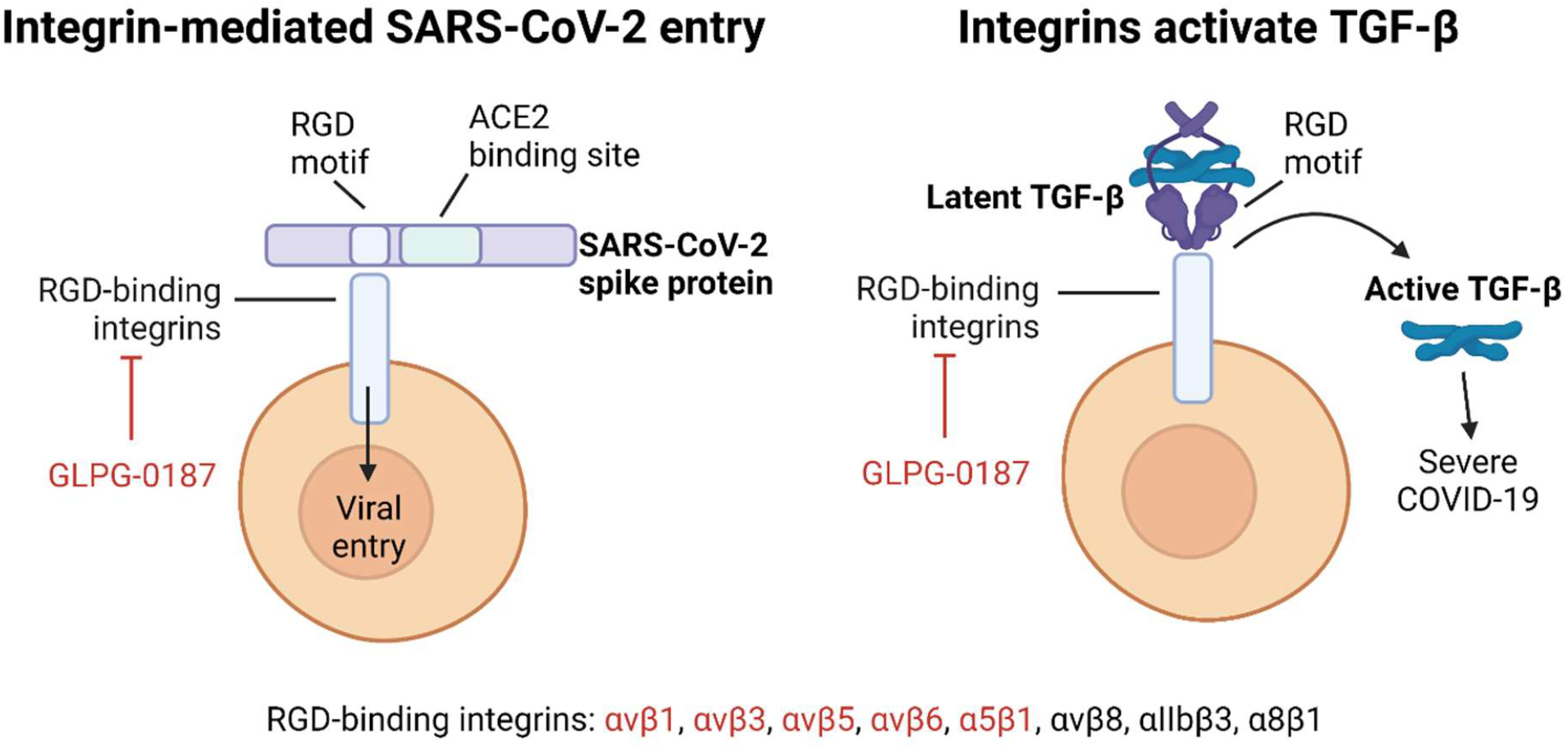
Integrins mediate SARS-CoV-2 infection and activation of TGF-β in human cells, facilitating viral pathogenesis. In addition to an ACE2-binding site, SARS-CoV-2 spike protein contains an RGD motif that binds RGD integrins, allowing viral infection (left). RDG-binding integrins are a major regulator of activation of latent TGF-β, which can be blocked via integrin inhibition (right). Active TGF-β may lead to more severe COVID-19 pathogenesis. GLPG-0187 is a pan-integrin inhibitor of the integrins (shown in red text) and has the potential to mitigate TGF-β mediated disease severity.

We previously demonstrated the feasibility of a SARS-CoV-2 pseudovirus model system to evaluate the effects of drug treatment on viral infection (14). Here, we show that the pan-integrin inhibitor GLPG-0187 inhibits infection of multiple pseudovirus variants in HSAE cells, including the highly transmissible Delta variant which was the most prevalent strain as of August 2021 (15) and the Omicron variant which became the most prevalent in December 2021 (www.who.int/news/item/26-11-2021-classification-of-omicron-(b.1.1.529)-sars-cov-2-variant-of-concern). This finding is clinically relevant as GLPG-0187 is in Phase I for treatment of solid tumors (16) and has shown a favorable toxicity profile in patients (17). GLPG-0187 targets the integrins αvβ1, αvβ3, αvβ5, αvβ6, and α5β1, which in addition to allowing infection of the virus may play a potential role in SARS-CoV-2 pathogenesis by mediating activation of TGF-β, angiogenesis, lung injury, and inflammation (18).

We previously demonstrated that treatment of HSAE cells with various MEK inhibitor (MEKi) compounds such as VS-6766 reduced cellular expression of ACE2 and inhibited pseudovirus infection (14). Thus, we hypothesized that GLPG-0187 and VS-6766 may have an additive or synergistic inhibitory effect on pseudovirus infection. VS-6766 received FDA Breakthrough Therapy Designation in combination with defactinib for treatment of ovarian cancer in 2021 (19), easing potential clinical translation.

We previously reported that SARS-CoV-2 pathogenesis can lead to myriad changes in cytokine, chemokine, and growth factor profiles in patient plasma samples, and that these changes are associated with disease severity (20). We recognized in our previous study that COVID-19 disease severity was associated with macrophage activation syndrome. Integrins can activate TGF-β, a growth factor secreted as a latent complex, which plays a role in immune response (21), fibrosis (22), and viral replication (23). The TGF-β complex consists of three proteins including TGF-β, latency-associated protein (LAP), and an extracellular matrix-binding protein. LAP contains an RDG integrin-binding site which mediates activation of latent TGF-β via RGD-binding integrins (24). The chronic immune response observed with SARS-CoV-2 is believed to be mediated by TGF-β (25). Thus, it has been suggested that SARS-CoV-2 pathogenesis could be controlled via modulation of TGF-β (26).

Our current study suggests that Omicron may less likely infect lower airway cells in the lung than other COVID-19 variants, and that integrin inhibitors have the potential to both prevent infection with SARS-CoV-2, including the Delta and Omicron variants, and to decrease TGF-β levels, resulting in a decrease in COVID-19 severity, hospitalization and death, especially in vulnerable and unvaccinated populations.

## Methods

### Cell culture

HSAE cells (ATCC PCS-301-010) were cultured in Airway Epithelial Cell Basal Medium (ATCC PCS-300-030) supplemented with the Bronchial Epithelial Cell Growth Kit (ATCC PCS-300-040) at 37°C in humidified atmosphere containing 5% CO2.

### SARS-CoV-2 pseudoviruses and cell entry assays

We developed a SARS-CoV-2 pseudovirus model system that uses pseudotyped SARS-CoV-2 viruses with a lentiviral core and a variety of SARS-CoV-2 spike protein variants on its envelope. To assess infectivity of normal human small airway epithelial (HSAE) cells, we used flow cytometry to quantify infected cells that express a fluorescence protein ZsGreen. A replication incompetent SARS-CoV-2 pseudovirus was generated using a lentiviral packaging system as previously described (14). Briefly, 293FT cells (Invitrogen) at 75% confluency were co-transfected with the backbone vector pHAGE-fullEF1α-Luciferase-IRES-ZsGreen, plasmids expressing lentiviral proteins Tat, Rev and Gag/Pol, and plasmids expressing D614 or D614G S protein (a gift from Dr. Hyeryun Choe, The Scripps Research Institute, Jupiter, FL), or S protein with N501Y, E484K, N501Y+E484K or N501Y+E484K+K417N mutations. An S protein expression plasmid construct containing all Beta variant (B.1.351) mutations and another S protein construct containing all Delta variant (B.1.617.2) mutations were gifts from Drs. Markus Hoffmann and Stefan Poehlmann (27) (German Primate Center, Goettingen, Germany). An S protein expression plasmid construct containing all Omicron variant (B.1.1.529) mutations (28) was custom-made by GenScript (Piscataway, NJ): pcDNA3.1(+)-SARS-CoV-2-Omicron-(6xHis)-Spike (human codon). The variant S genes in the above constructs were sequenced to confirm all the corresponding mutations (**Table 1**). A plasmid expressing VsVg protein instead of the S protein was used to generate a pantropic control lentivirus. Cell culture supernatants were collected, filtered, concentrated using ultra-centrifugation, aliquoted, and frozen at –80°C. Virus titer was determined using Lenti-X(tm) p24 Rapid Titration ELISA Kit (TaKaRa) and lentiviral particles were analyzed on an SDS-PAGE gel followed by Western blot to detect C-terminal FLAG-tagged S protein. HSAE cells were pre-treated with the integrin inhibitor GLPG-0187 (Galapagos NV, Mechelen, Belgium), MEK inhibitor VS-6766 (Verastem Oncology, Needham, MA), or both, for 2-27 hours. Following drug treatment, HSAE cells were spin-infected with SARS-CoV-2 pseudoviruses or a pantropic VsVg positive control lentivirus in a 12-well plate (931 g, 2 hours, 30°C with 8 μg/ml polybrene). Analysis of ZsGreen+ cells was conducted by flow cytometry 20-24 hours after infection using a BD LSRII flow cytometer and FlowJo software.

**Table 1.** Description of several SARS-CoV-2 viral spike variants that are represented in this study with experimental pseudoviruses.

| Variant | Description |
| --- | --- |
| Omicron (B.1.1.529) | Dominant strain as of December 2021. Spike mutations include: A67V, Δ69-70, T95I, G142D, Δ143-145, Δ211, L212I, ins214EPE, G339D, S371L, S373P, S375F, K417N, N440K, G446S, S477N, T478K, E484A, Q493R, G496S, Q498R, N501Y, Y505H, T547K, D614G, H655Y, N679K, P681H, N764K, D796Y, N856K, Q954H, N969K, L981F. |
| Delta (B.1.617.2) | Dominant strain as of August 2021. Spike mutations include: T19R, G142D, E156G, F157Δ, R158Δ, L452R, T478K. D614G, P681R, D950N. |
| Beta (B.1.351) | Prevalent in late 2020. Spike mutations include: L18F, D80A, D215G, Δ242-244, R246I, K417N, E484K, N501Y, D614G, A701V . |
| D614G | Dominant strain in the spring of 2020 |
| D614 | Prevalent strain in early 2020 |
| N501Y | A common mutation in the Alpha (B.1.1.7), Beta (B.1.351), and Gamma (P.1) variants |
| E484K | A common mutation in the Beta (B.1.351) and Gamma (P.1) variants |
| N+E (N501Y + E484K) | Common mutations in the Beta (B.1.351) and Gamma (P.1) variants |
| NEK (N501Y + E484K + K417N) | Common mutations in the Beta (B.1.351) and Gamma (P.1) variants |
| R785A | Furin-cleavage site mutated |

### Human plasma samples

COVID-19 (+) human plasma samples were received from the Lifespan Brown COVID-19 Biobank at Rhode Island Hospital (Providence, RI, USA). All patient samples were deidentified but contained associated clinical information, as described. The IRB study protocol “Pilot Study Evaluating Cytokine Profiles in COVID-19 Patient Samples” did not meet the definition of human subjects research by either the Brown University or the Rhode Island Hospital IRBs.

### IRB/oversight of exemption for the research (as previously described) (29)

COVID-19 (+) and (-) human plasma samples were received from the Lifespan Brown COVID-19 Biobank from Brown University at Rhode Island Hospital (Providence, Rhode Island). All patient samples were deidentified but included the available clinical information as described. The IRB study protocol Pilot Study Evaluating Cytokine Profiles in COVID-19 Patient Samples did not meet the definition of human subjects research by either the Brown University or the Rhode Island Hospital IRBs. This is based on the fact that the project used deidentified specimens from a biobank with a determination that this project did not meet the definition of human subjects research based on specific criteria as described below. The original samples were collected at Rhode Island hospital by the Lifespan Brown COVID-19 Biobank through an IRB-approved protocol that involved informed consent that was used by the biobank. We completed a human subjects determination form for the Human Subjects Protection Program at Brown University. We explained the purpose of our research and that we would be receiving deidentified samples from the COVID-19 biobank. We further answered questions about our study that led to the determination that our study constitutes research because we answered “yes” to the following two questions: 1-Does your proposed project involve a systematic investigation: that is a prospective plan that incorporates qualitative or quantitative data collection, and data analysis to answer a question?, and 2-Is the intent of your proposed project to develop or contribute to generalizable knowledge; that is to create knowledge from which conclusions will be drawn that can be applied to populations beyond the specific population from which it was collected. In addition, we answered “no” to four questions regarding whether the project involves human subjects. The questions were 1-Does your proposed project involve an intervention: that is a physical procedure or manipulation or a living individual (or their environment) to obtain information about them? 2-Does your proposed project involve an interaction; that is communication or contact with a living individual (in person, online, or by phone) to obtain information about them? 3-Does your proposed project involve identifiable private information or identifiable biospecimen; that is receipt or collection of private information or biospecimen about a living individual to obtain information about them? and 4-Does your proposed project involve coded information/ biospecimens; that is where a link exists that could allow information about a living individual to be reidentified AND you are able to access the link? Since we answered “no” to all these questions, our proposed project did NOT involve “Human Subjects.” Based on the information included in the Human Subjects Determination Form, The Human Research Protection Program at Brown University agreed with the investigator’s self-determination that the project does not meet the definition of human subjects research. This determination was made by the Human Research Protection Program at Brown University on June 17, 2020.”

### Cytokine profiling

A Human Magnetic Luminex Performance Assay TGF-β1 Base Kit (Cat # LTGM100, R&D Systems, Inc., Minneapolis, MN) was run on a Luminex 200 Instrument (LX200-XPON-RUO, Luminex Corporation, Austin, TX) according to the manufacturer’s instructions. Total TGF-β was quantified by activating patient samples with 1N HCl, neutralizing with 1.2N NaOH/0.5M HEPES, and then immediately assaying for TGF-β1. Active TGF-β was quantified without sample activation or neutralization prior to analysis.

### Statistical Analysis

A spearman’s correlation was used to calculate statistical significance of scatter plots while the statistical significance between groups was determined using a One-way Anova followed by a post-hoc Tukey’s multiple comparisons test. A two-tailed, unpaired student’s t test was used to calculate statistical significance of pairs. The minimal level of significance was P < 0.05. Following symbols * and ** represent, P < 0.05 and P < 0.01, respectively.

## Results

### Integrin inhibition decreases infection of SARS-CoV-2 pseudovirus variants in human small airway epithelial cells

To test the inhibition of SARS-CoV-2 pseudovirus infection with the integrin inhibitor GLPG-0187, HSAE cells were pre-treated with 20 nM, 100 nM, 200 nM, or 1 µM GLPG-0187 for 2 hours followed by spin-infection with either a pseudovirus expressing the D614G spike protein variant or a VsVg positive control for 24 hours. Few ZsGreen+ cells were seen in the cells not treated with spin-infection, efficient viral infection was observed in cells treated with spin infection, and no effect of the inhibitor was observed on the VsVg positive control, as expected. Treatment with GLPG-0187 inhibited pseudovirus infection in a dose-dependent manner in the D614G variant **(Figure 2A)**. In addition to D614G, several other SARS-CoV-2 pseudovirus variants were also tested including D614, N501Y, E484K, N501Y + E484K (N+E), N501Y + E484K + K417N (NEK), R685A. Descriptions of these variants can be found in **Table 1**. HSAE cells pre-treated with 1 µM GLPG-0187 for 3 hours followed by spin-infection with pseudovirus variants for 20 hours demonstrated inhibition of viral infection of each variant **(Figure 2B)**. To test inhibition of Beta and Delta variant pseudovirus infection, HSAE cells were pre-treated with 1 or 2 µM GLPG-0187 for 2 hours followed by spin-infection with pseudovirus. Pre-treatment with the integrin inhibitor resulted in the most significant decrease in pseudovirus infection by the Delta variant **(Figure 2C)**. We conducted experiments with Omicron pseudovirus infection on HSAE cells with or without the integrin inhibitor GLPG-0187 **(Figures 2D and E)**. The results suggest that Omicron pseudovirus was less capable of infecting the small airway epithelial cells than D614G or Delta variant pseudovirus, which is in agreement with a recent study by Meng *et al*. (30). Nevertheless, the integrin inhibitor GLPG-0187 effectively blocked D614G, Delta and Omicron pseudovirus infection of HSAE cells.

**Figure 2.**
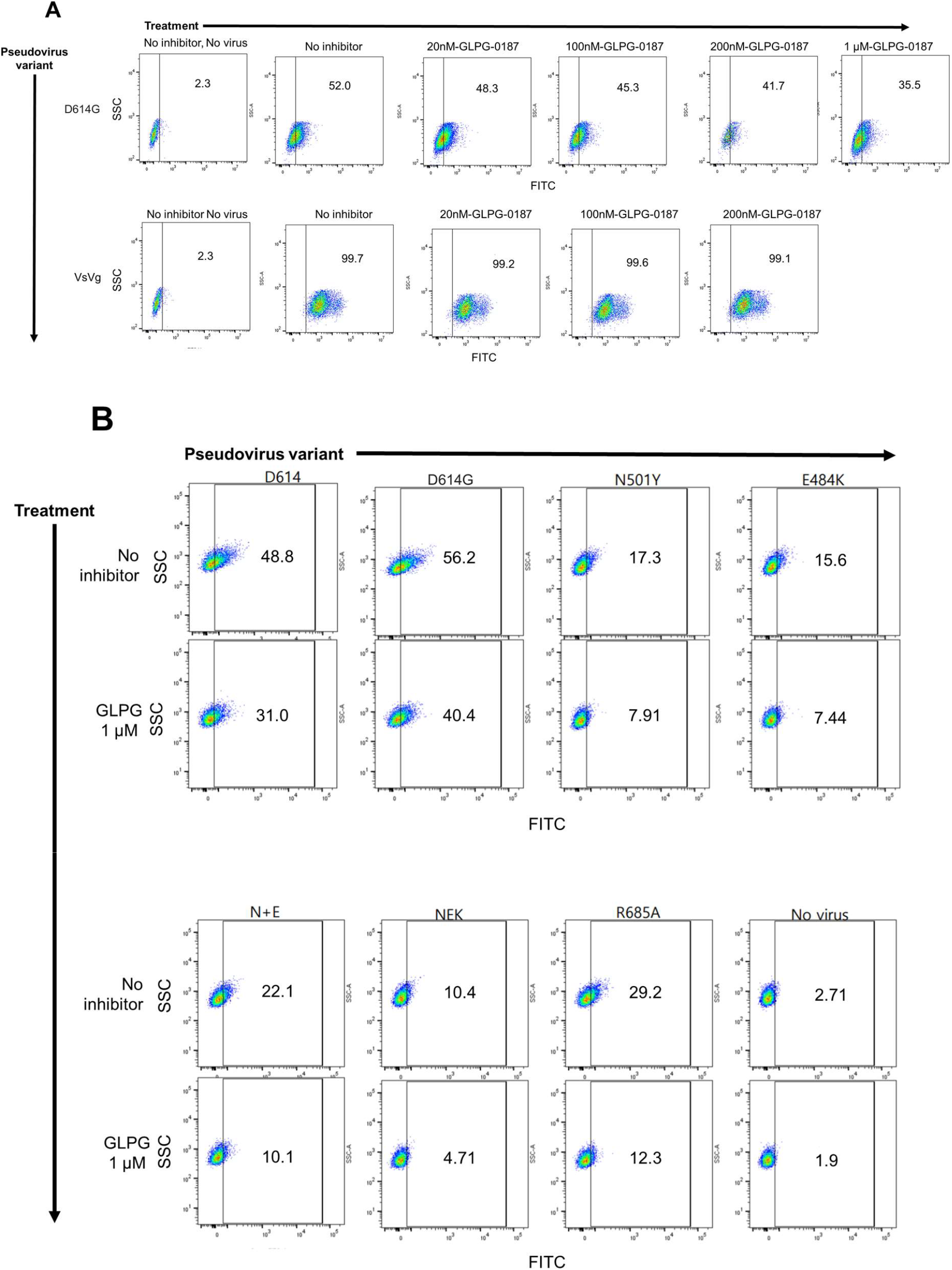
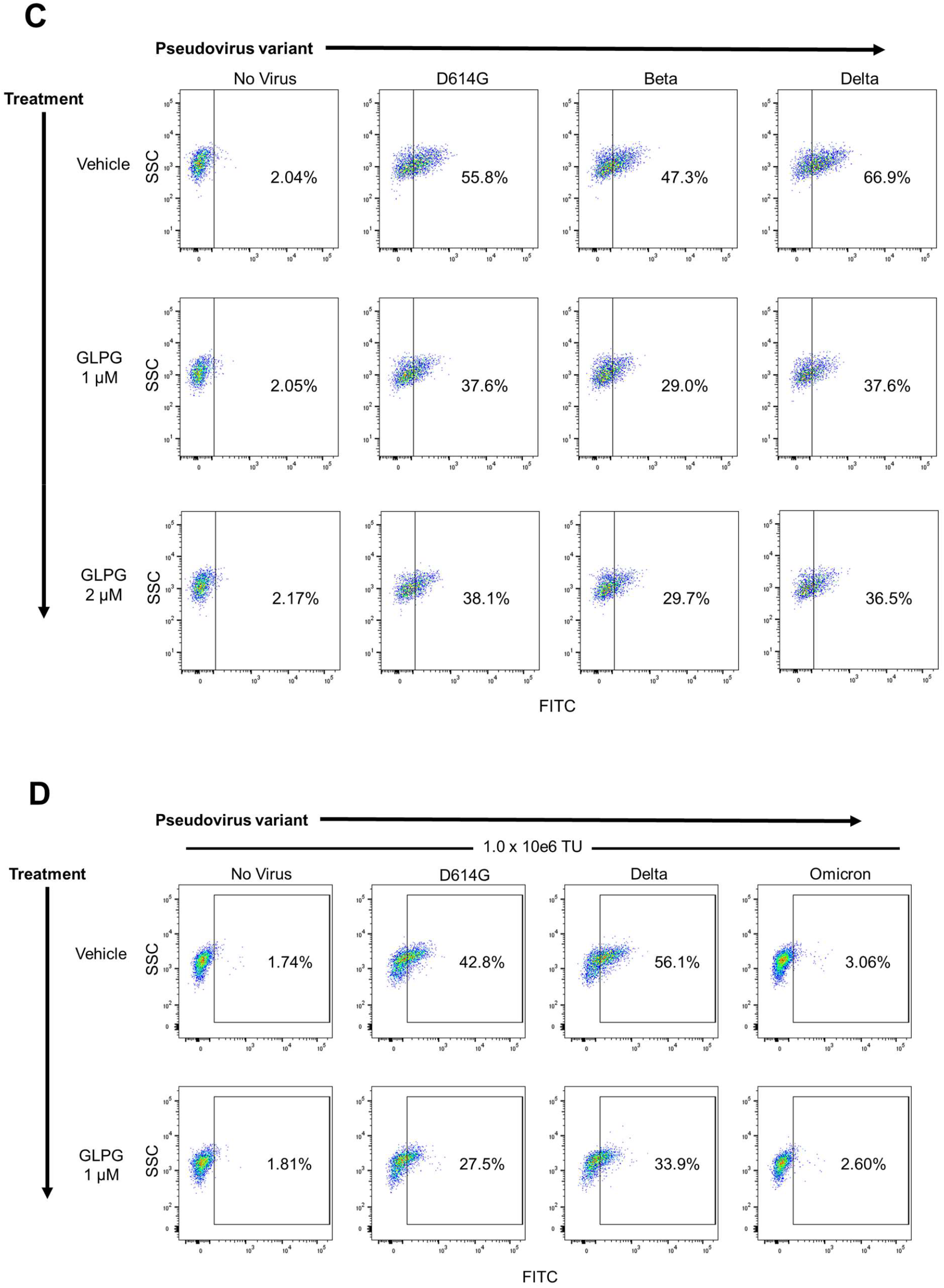
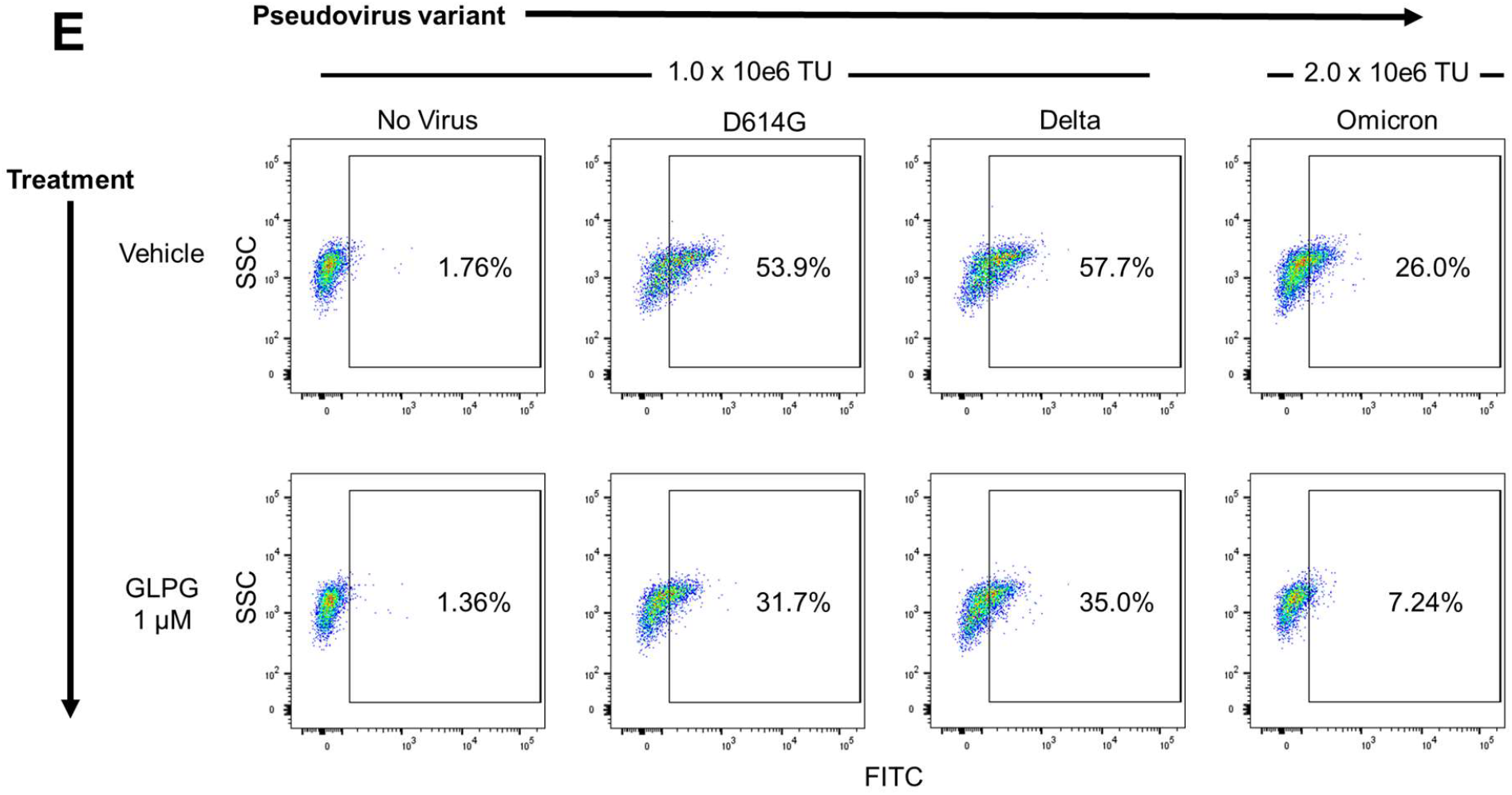
GLPG-0187 inhibits infection of SARS-CoV-2 pseudovirus variants D614, D614G, N501Y, E484K, N+E, NEK, R685A, Beta, Delta, and Omicron in small airway epithelial cells. (A) Treatment with 20 nM, 100 nM, 200 nM, or 1 µM GLPG-0187 for 2 hours inhibits infection by the D614G pseudovirus variant (24 hour infection time) in small airway epithelial cells compared to the VsVg positive control in a dose-dependent manner. (B) Treatment with 1 µM GLPG-0187 for 3 hours inhibits infection by the D614, D614G, N501Y, E484K, N+E, NEK, R685A pseudovirus variants (20 hour infection time). (C) Treatment with 1 µM or 2 µM GLPG-0187 for 2 hours inhibits infection by the D614G, Beta, and Delta pseudovirus variants (20 hour infection time). (D) Differential rates of infectivity across D614G, Delta, and Omicron variants observed after cells were spin-infected with the same amount of pseudovirus particles (1.0 × 10e6 transduction units (TU) per 1 × 10e5 cells/well). (E) Treatment with 1 µM GLPG-0187 for 2 hours inhibits infection by the Omicron pseudovirus variant (26 hour infection time).

### MEK inhibitor pre-treatment enhances inhibition of pseudovirus infection by GLPG-0187 in human small airway epithelial cells

We have previously demonstrated that MEKi compounds including VS-6766 reduce cellular expression of ACE2 and inhibit pseudovirus infection of multiple human cell types (14). Thus, we hypothesized that VS-6766 and GLPG-0187 could have an additive or synergistic inhibitory effect on pseudovirus infection of lung epithelial cells. To investigate this, we pre-treated HSAE cells with either 5 µM VS-6766 for 24 hours, 1 µM GLPG-0187 for 3 hours, or 5 µM VS-6766 for 24 hours followed by an additional 3 hours with GLPG-0187. After drug treatment, cells were spin-infected with the D614G pseudovirus for 20 hours. As expected, VS-6766 and GLPG-0187 single treatment inhibited pseudovirus infection when compared to the positive control. Combination treatment enhanced the inhibition of pseudovirus infection compared to single agent treatment with either VS-6766 or GLPG-0187 **(Figure 3)**.

**Figure 3.**
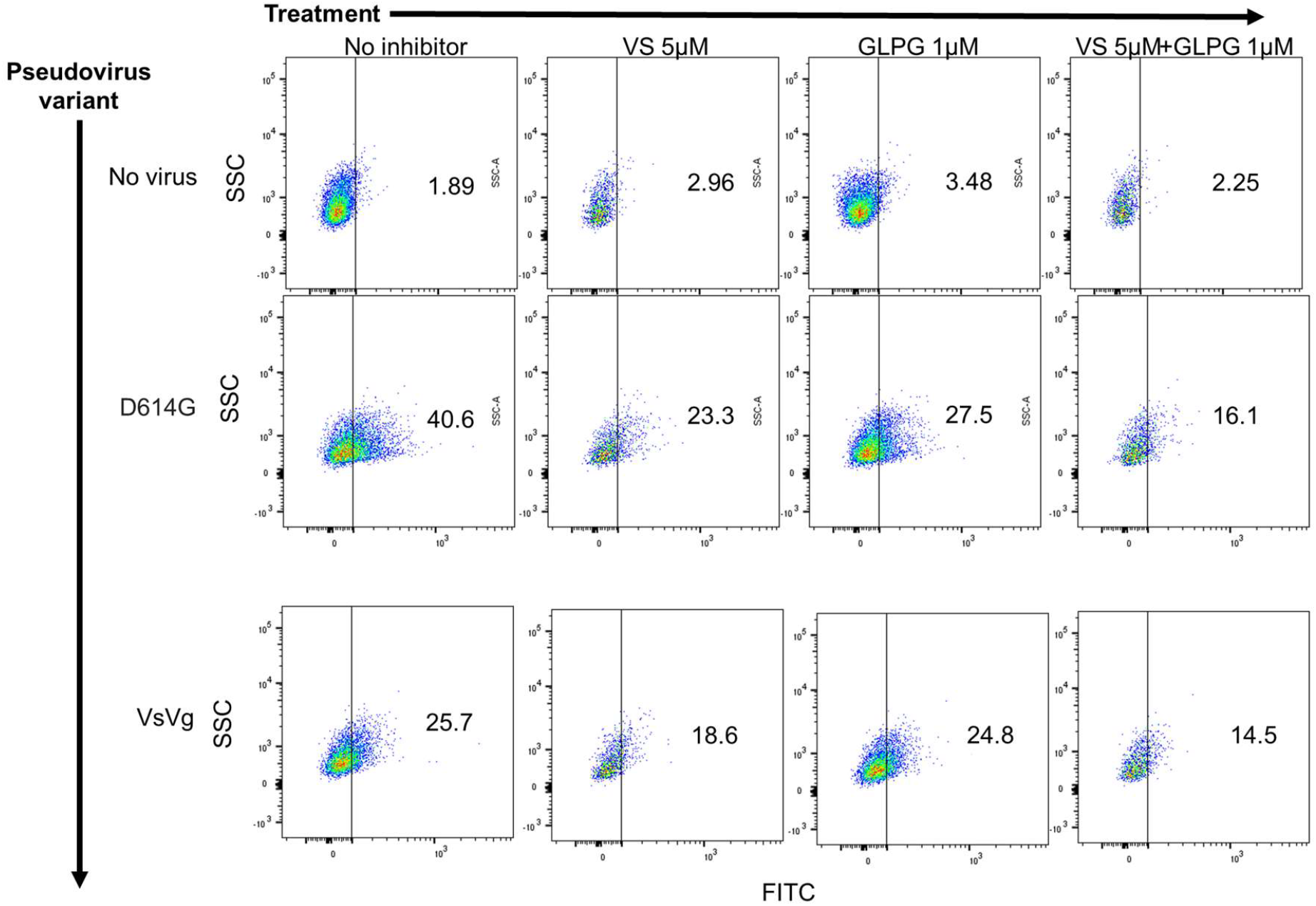
MEK inhibitor VS-6766 enhances the inhibition of SARS-CoV-2 pseudovirus infection by integrin inhibitor GLPG-0187 in small airway epithelial cells. Treatment with 5 µM VS-6766 for 24 hours or with 1 µM GLPG-0187 for 3 hours inhibits infection by the D614G pseudovirus variant (20 hour infection time) in small airway epithelial cells compared to the VsVg positive control. Combination treatment involved 24 hour pre-treatment with VS-6766 followed by an additional 3 hours of treatment with GLPG-0187.

### Plasma TGF-β1 levels correlate with age, race, and number of medications administered upon presentation with COVID, but not with sex

Because it has been previously shown that the chronic immune response observed with SARS-CoV-2 is mediated by TGF-β, we sought to compare the levels of TGF-β1 in plasma samples from COVID (+) patients upon admission to the emergency department (ED). We chose to focus on TGF-β1, as opposed to TGF-β2 and 3, since it had been previously shown that SARS-CoV-2 infection increased TGF-β1 expression in human epithelial cells and was a driver of lung fibrosis (31). We analyzed the levels of total TGF-β1 in COVID (+) plasma samples and found a significant correlation between TGF-β1 concentration (pg/mL) and age **(Figure 4A)**. We also found significant variations in TGF-β1 concentrations depending on the patient’s self-reported race or ethnicity, with notably higher levels of the growth factor in White and Hispanic or Latino populations, and notably lower levels in Black and Asian or Pacific Islander populations **(Figure 4B)**. We next grouped patients by the number of medications they received upon disease presentation to the emergency department **(Figure 4C)**. Medications reported included ibuprofen, acetaminophen, bronchodilators (e.g., Albuterol), steroids (e.g., Prednisone), azithromycin, hydroxychloroquine, antibiotic, or other. We noticed statistically significant decreased plasma TGF-β1 concentrations in patients that received 2-4 medications in the Emergency Department (ED), as compared to patients who received 0-1. Next, we grouped patients by number of symptoms self-reported upon admission to the ED and noted a positive trend between TGF-β1 levels and the number of symptoms, although not significant **(Figure 4D)**. When comparing TGF-β1 levels between male and female, we did not note a significant difference **(Figure 4E)**.

**Figure 4.**
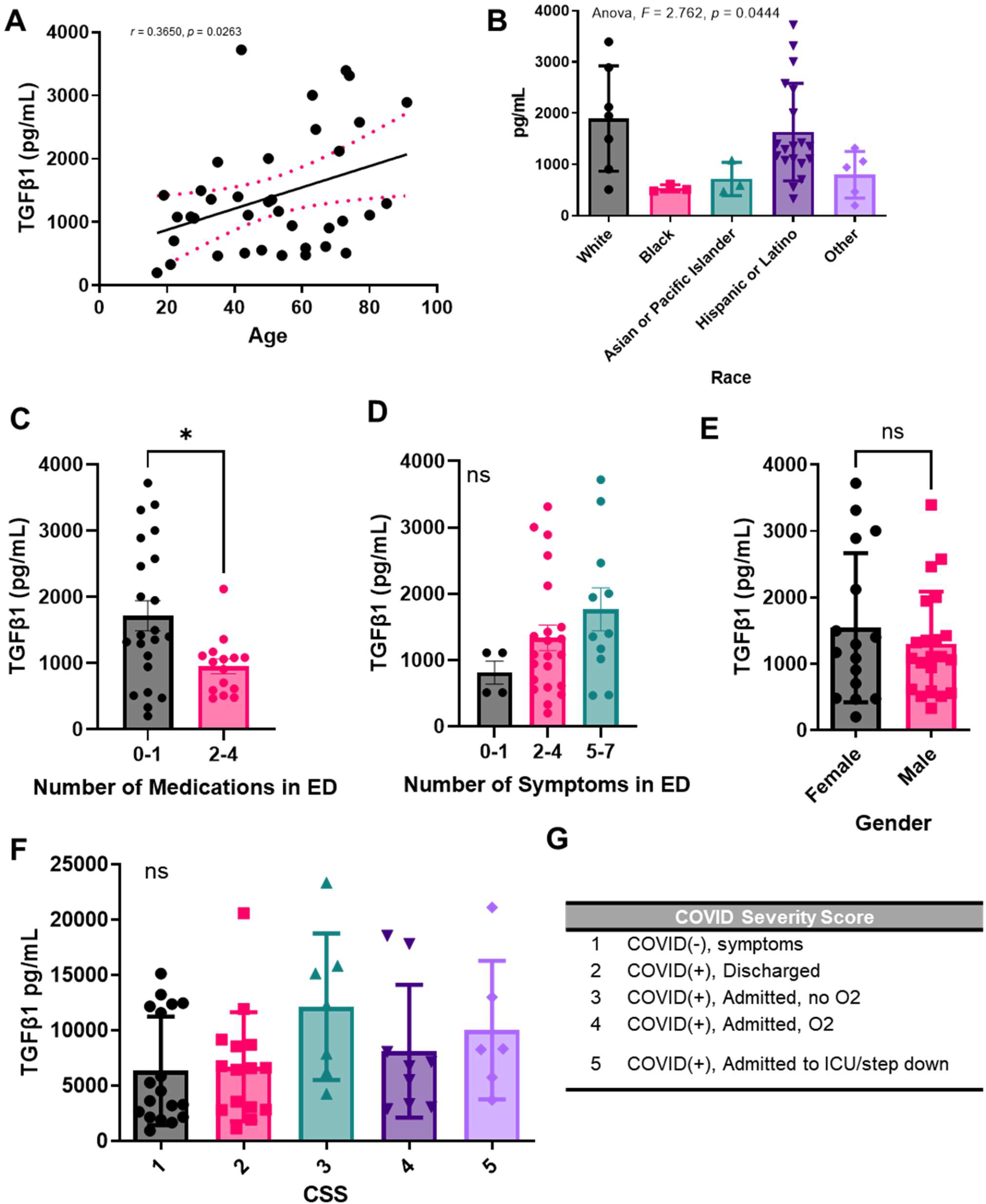
Plasma TGF-β1 levels correlate with age, race, and number of mediations administered upon presentation with COVID to the ED, but not with sex. Total TGF-β1 levels were detected in activated plasma samples. TGF-β1 plasma concentration correlation with (A) age, (B) race, (C) number of mediations administered upon presentation with COVID-19 to the emergency department (ED), (D) number of symptoms reported upon presentation to the ED, (E) sex, or (G) COVID-19 severity score. (G) COVID-19 severity score (CSS) legend. Sample values are reported in pg/mL (n=81 samples).

We also compared TGF-β1 levels in patients based on our COVID-19 Severity Score (CSS) (**Figure 4F)** which was based on the presence or absence of symptoms, patient oxygen requirements, and whether or not the patient was admitted to the ICU/step down units **(Figure 4G)**. We again noted a positive trend between growth factor levels and increasing COVID severity. Because we were interested in the role of TGF-β1 in the pathogenesis of other diseases as well, we also compared TGF-β1 levels in patients with a prior history of disease including chronic lung disease, chronic kidney disease, chronic heart disease, pneumonia, high blood pressure, diabetes, previous strike, and abnormal chest x-ray upon ED admission **(Supplementary Figure 1)**. However, due to a limited sample size, we only noted a significant increase in TGF-β1 in the patients with a history of chronic kidney disease as compared to those without a history of chronic kidney disease. Others have reported that treatment of breast cancer cells with GLPG-0187 decreased TGF-β signaling (32). Thus, treatment with GLPG-0187 may especially benefit populations of patients with high levels of TGF-β1.

### Active plasma TGF- β1 levels correlate with total TGF-β1 levels

Because we were interested in the concentrations of both active TGF-β1 and total TGF-β1, we next analyzed the patient plasma samples for active TGF-β1. We observed similar trends as described above and noted that active plasma TGF-β1 levels correlate with total TGF-β1 levels **(Figure 5)**. We again noted a significant correlation between TGF-β1 plasma concentration and patient age **(Figure 5A)**. We similarly noted higher levels of the growth factor in self-reported White and Hispanic or Latino populations, and notably lower levels in Black and Asian or Pacific Islander populations **(Figure 5B)**. When comparing active TGF-β1 levels between sexes, we again did not note a significant difference **(Figure 5C)**. Finally, we again noted a positive trend between active TGF-β levels and increasing COVID severity, as determined by our CSS criteria **(Figure 5D)**.

**Figure 5.**
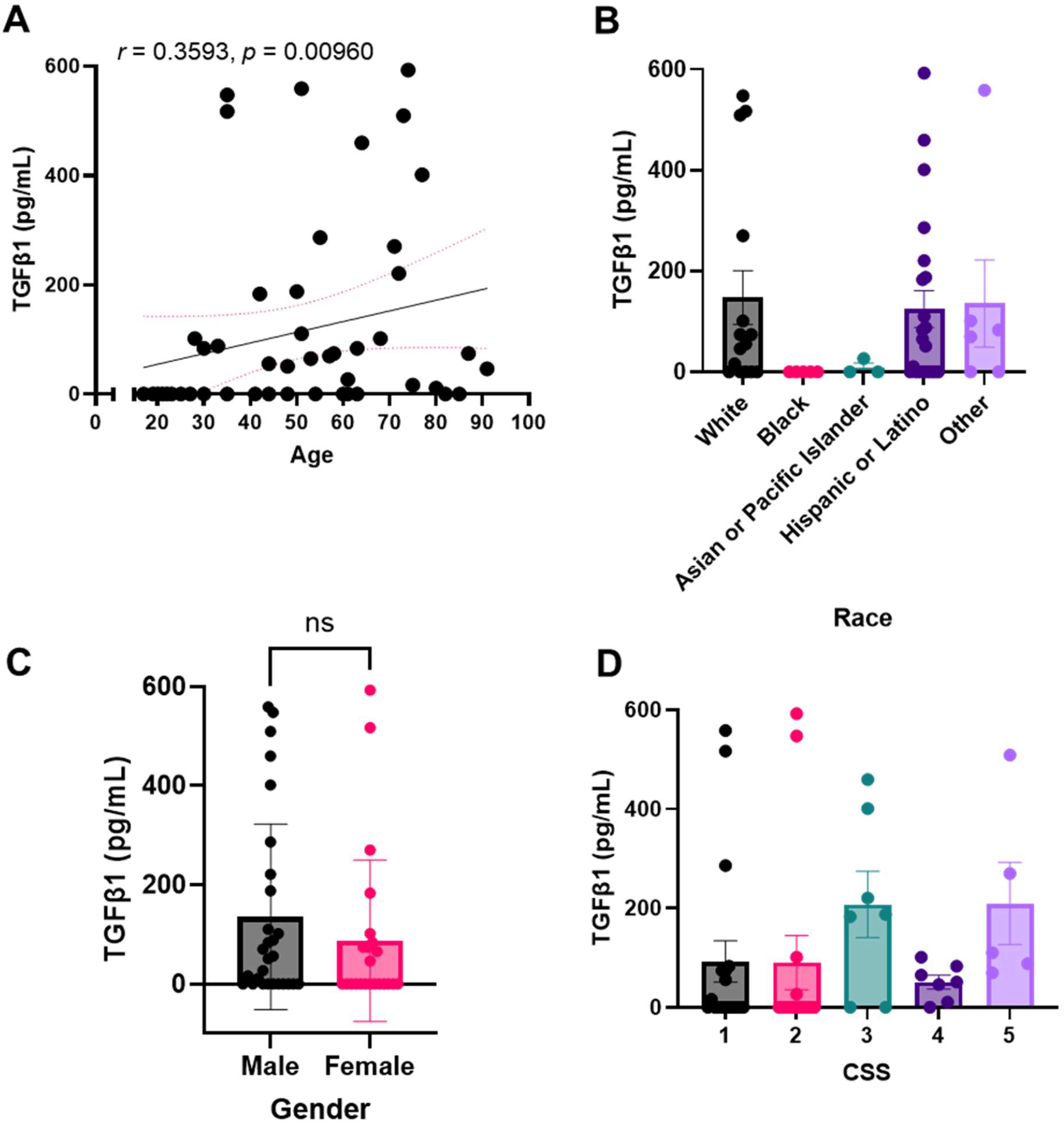
Active plasma TGF-β1 levels correlate with total TGF-β1 levels. Active TGF-β1 levels were detected in non-activated plasma samples. TGF-β1 plasma concentration correlation with (A) age, (B) race, (C) sex, or (D) COVID-19 severity score (CSS). Sample values are reported in pg/mL (n=81 samples).

## Discussion

SARS-CoV-2 remains a significant challenge in global health and new treatment options are needed, especially for vulnerable and unvaccinated populations. As of November 2021, the Delta variant accounted for more than 99% of COVID-19 cases and infection with this variant may result in an increased likelihood of hospitalization (33). Since then, the Omicron variant rapidly spread around the globe and became the dominant strain in many parts of the world (www.who.int/news/item/26-11-2021-classification-of-omicron-(b.1.1.529)-sars-cov-2-variant-of-concern). Our current study suggests that integrin inhibition inhibits infection of multiple SARS-CoV-2 pseudovirus variants in HSAE cells, and that GLPG-0187 may be particularly effective in inhibiting infection by the Delta variant. As HSAE cells are thought to have very low expression of ACE2, alternative targets such as RGD-binding integrins may have particular value for treatment of COVID-19 (14, 34, 35). Our findings also suggest that combination treatment with a MEKi enhances this effect. In addition to inhibiting viral infection, it is possible that integrin inhibition could provide benefit to COVID-19 patients by reducing levels of active TGF-β, as integrins are a major regulator of TGF-β activation (36). Limited prior studies have reported that COVID-19 patients may have higher levels of TGF-β compared to healthy controls, which may mediate some of the complications in severe COVID-19 patients (18).

Thus, benefit from GLPG-0187 -/+ MEKi treatment in COVID-19 patients may be mediated through (1) inhibition of viral infection and (2) inhibition of TGF-β activation. Integrin inhibition may especially provide benefit to COVID-19 patient populations with particularly high levels of TGF-β such as elderly, White and Hispanic or Latino patients and patients who receive few medications in the ED, report a high number of symptoms, have a high CSS, and/or have a history of chronic kidney disease. Two limitations of this study include the small sample size of plasma samples from patients with COVID-19, as well as the lack of serial samples over time from the same patient. Because we only analyzed plasma from patients upon admission to the emergency department, we may have missed fluctuations in TGF-β concentrations, which are thought to peak during the first two weeks post-infection in severe COVID-19 cases (37). Future work should monitor levels of TGF-β in serial patient samples.

Since its first identification in South Africa in November, 2021, the SARS-CoV-2 Omicron variant raised serious concerns of a significant reduction in efficacy of vaccines and monoclonal antibody treatments and an increased risk of reinfection due to numerous mutations in its spike protein, which is the antigenic target of infection- and vaccine-elicited antibodies against SARS-CoV-2. Currently, the Omicron variant is on track to outcompete the Delta variant as cases have soared to record highs in parts of Europe and now the U.S. according to the data released by Johns Hopkins University (https://coronavirus.jhu.edu/data). A number of recent studies suggest that much of the Omicron variant’s dominance comes down to its ability to evade the body’s immune defenses (28, 30, 38-40). However, earlier analyses of patients in South Africa suggest Omicron-infected individuals had a reduced risk of severe disease when compared to Delta-infected individuals (39). In the first findings on how the Omicron variant infects the respiratory tract, researchers from Hong Kong University reported that the virus multiplies 70 times faster in the bronchi than Delta and the original SARS-CoV-2 virus (https://www.med.hku.hk/en/news/press/20211215-omicron-sars-cov-2-infection). In a potential clue regarding lower disease severity, they found that Omicron replication was less efficient in deeper lung tissue, more than 10 times lower than the original virus. In a recent study by Meng et al. (30), the investigators found that despite three mutations predicted to favor spike S1/S2 cleavage, observed cleavage efficiency is substantially lower than for Delta, and Omicron pseudovirus entry into lower airway organoids and Calu-3 lung cells was thus impaired. In a latest study on mice and hamsters (40), Omicron produced less-damaging infections, often limited largely to the upper airway: the nose, throat and windpipe. The variant did much less harm to the lungs, where previous variants would often cause scarring and serious breathing difficulty. In our current study, we found that the Omicron pseudovirus was less capable of infecting the small airway epithelial cells than D614G or Delta variant pseudovirus, and that integrin inhibition effectively blocked D614G, Delta and Omicron pseudovirus infection of the small airway epithelial cells. Combined, these observations highlight that Omicron has gained immune evasion properties whilst compromising cell entry in lung cells, with possible implications for altered pathogenicity. In addition, targeting alternative viral infection routes such as integrin-mediated cell entry and dampening TGF-β1-mediated disease severity may have therapeutic implications, especially for vulnerable and unvaccinated populations.

It is possible that GLPG-0187 inhibits pseudovirus variant infection by an off-target effect on ACE2. It is also possible that (1) the virus may infect ACE2 negative cells by using RGD-binding integrins as an alternative receptor to ACE2 and/or (2) the RGD motif functions as a coreceptor that enhances viral infection via ACE2. Future work should involve similar experiments in cells that are completely ACE2 negative either naturally or by genetic modification, as low levels of ACE2 expression may still be relevant for viral infection. Future work may also include development of a pseudovirus with a mutated RGD motif to assess effects on viral infection as well as to analyze which RGD-integrin(s) are important for SARS-CoV-2 infection. Our results nonetheless demonstrate that GLPG-0187 inhibits pseudovirus entry, providing rationale for further investigation of integrin inhibitors as a potential therapy for COVID-19. Moreover, our findings offer a combinatorial strategy combining an integrin inhibitor with a MEK inhibitor as a therapeutic strategy against COVID-19 including Delta and Omicron variants. These strategies could be further tested in clinical trials with particularly at risk patients with COVID-19 infection who are unvaccinated, immunosuppressed, or with risk factors such as comorbidities including cancer.

## Data Availability

All data produced in the present work are contained in the manuscript

## Acknowledgements

The work was supported by a Brown University COVID-19 Seed Grant (to W.S.E-D.), and the Mencoff Family Professorship at Brown University (W.S.E-D.). W.S.E-D. is an American Cancer Society Research Professor. O.L. was supported in part by a grant from the National Institutes of Health (P20 GM119943). The COVID-19 Biobank through which plasma samples were obtained was supported by Institutional Development Award Number U54GM115677 from the National Institute of General Medical Sciences of the National Institutes of Health, which funds Advance Clinical and Translational Research (Advance-CTR). E-Y.S. was supported in part by several grants, from the National Institutes of General Medical Science (5P30GM122732-05), The Rhode Island Foundation (841-20210959), the University of Rhode Island/NIGMS (0009351/10262021), and Brown Physicians Incorporated (BPI Research Award). K.H. and L.C. were supported by the Teymour Alireza P’98, P’00 Family Cancer Research Fund established by the Alireza Family. The content is solely the responsibility of the authors and does not necessarily represent the official views of the National Institutes of Health. Figures were created with BioRender.com.

## Author Disclosures

None of the co-authors have disclosed relationships that are relevant for this work.

---

**Supplementary Figure 1.**
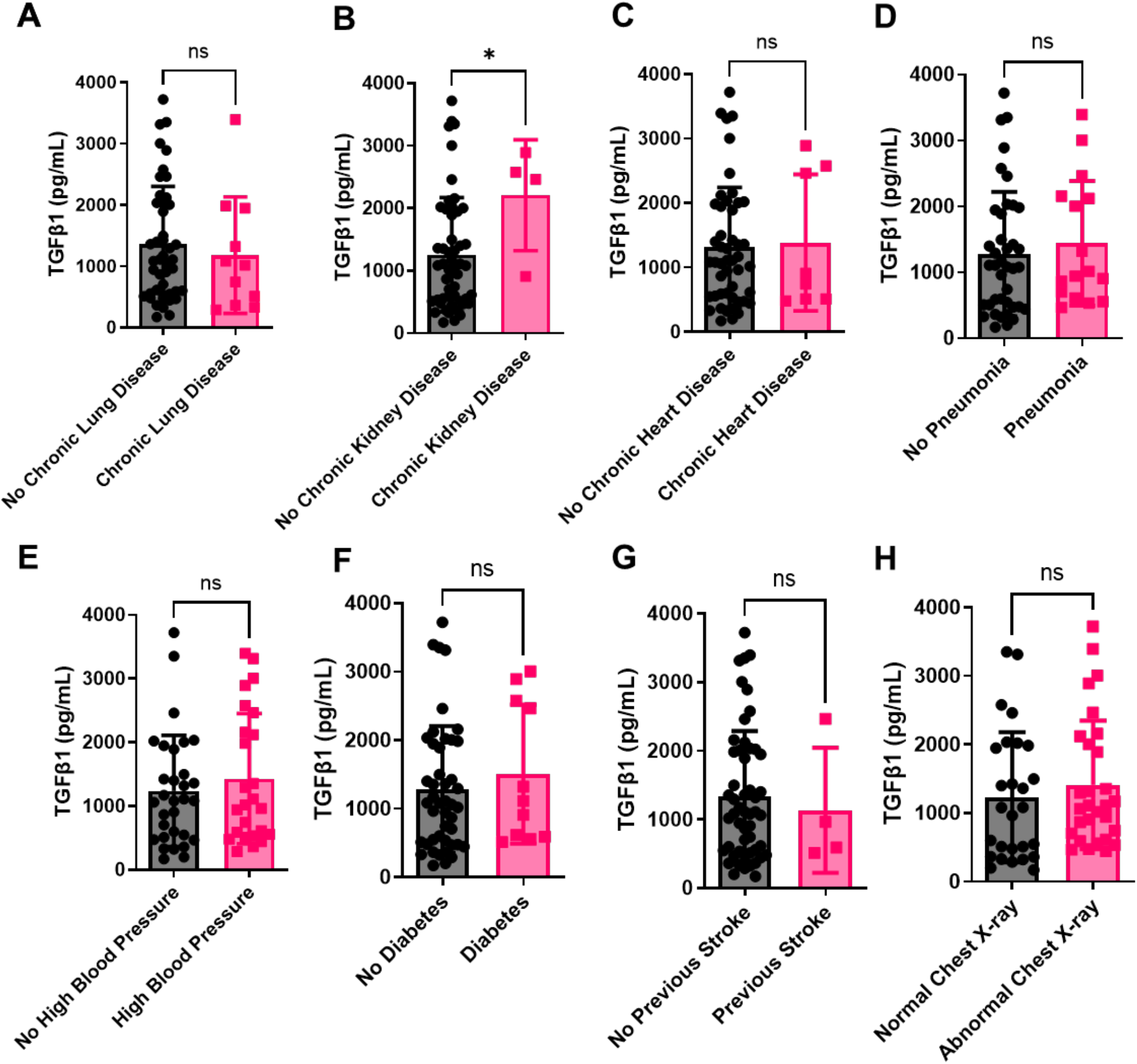
Plasma TGF-β1 levels are elevated in patients with a history of kidney disease. Total TGF-β1 levels were detected in activated plasma samples. TGF-β1 plasma concentration correlation with a history of (A) chronic lung disease, (B) chronic kidney disease, (C) chronic heart disease (D) pneumonia upon presentation to the emergency department (ED), (E) high blood pressure, (F) diabetes, (G) stroke, or (H) chest x-ray upon presentation to the ED. Sample values are reported in pg/mL (n=81 samples).

## Notes

### Competing Interest Statement

The authors have declared no competing interest.

### Funding Statement

This study was funded by a Brown University COVID-19 Seed Grant and by a grant from the National Institutes of Health (P20 GM119943) The COVID-19 Biobank through which plasma samples were obtained was supported by Institutional Development Award Number U54GM115677 from the National Institute of General Medical Sciences of the National Institutes of Health E-YS was supported in part by grants from the National Institutes of General Medical Science (5P30GM122732-05) The Rhode Island Foundation (841-20210959) The University of Rhode Island/NIGMS (0009351/10262021) and Brown Physicians Incorporated (BPI Research Award) KEH and LC were supported by the Teymour Alireza P98 P00 Family Cancer Research Fund established by the Alireza Family.

### Author Declarations

Ethics committee/IRB of Brown University waived ethical approval for this work. COVID-19 (+) and (−) human plasma samples were received from the Lifespan Brown COVID-19 Biobank from Brown University at Rhode Island Hospital (Providence, Rhode Island). All patient samples were deidentified but included the available clinical information as described. The IRB study protocol Pilot Study Evaluating Cytokine Profiles in COVID-19 Patient Samples did not meet the definition of human subjects research by either the Brown University or the Rhode Island Hospital IRBs. This is based on the fact that the project used deidentified specimens from a biobank with a determination that this project did not meet the definition of human subjects research based on specific criteria as described below. The original samples were collected at Rhode Island hospital by the Lifespan Brown COVID-19 Biobank through an IRB-approved protocol that involved informed consent that was used by the biobank. We completed a human subjects determination form for the Human Subjects Protection Program at Brown University. We explained the purpose of our research and that we would be receiving deidentified samples from the COVID-19 biobank. We further answered questions about our study that led to the determination that our study constitutes research because we answered yes to the following two questions: 1- Does your proposed project involve a systematic investigation: that is a prospective plan that incorporates qualitative or quantitative data collection, and data analysis to answer a question?, and 2- Is the intent of your proposed project to develop or contribute to generalizable knowledge; that is to create knowledge from which conclusions will be drawn that can be applied to populations beyond the specific population from which it was collected. In addition, we answered no to four questions regarding whether the project involves human subjects. The questions were 1- Does your proposed project involve an intervention: that is a physical procedure or manipulation or a living individual (or their environment) to obtain information about them? 2- Does your proposed project involve an interaction; that is communication or contact with a living individual (in person, online, or by phone) to obtain information about them? 3-Does your proposed project involve identifiable private information or identifiable biospecimen; that is receipt or collection of private information or biospecimen about a living individual to obtain information about them? and 4- Does your proposed project involve coded information/ biospecimens; that is where a link exists that could allow information about a living individual to be reidentified AND you are able to access the link? Since we answered no to all these questions, our proposed project did NOT involve Human Subjects. Based on the information included in the Human Subjects Determination Form, The Human Research Protection Program at Brown University agreed with the investigator's self-determination that the project does not meet the definition of human subjects research. This determination was made by the Human Research Protection Program at Brown University on June 17, 2020.

